# Trialstreamer: a living, automatically updated database of clinical trial reports

**DOI:** 10.1101/2020.05.15.20103044

**Authors:** Iain J Marshall, Benjamin Nye, Joël Kuiper, Anna Noel-Storr, Rachel Marshall, Rory Maclean, Frank Soboczenski, Ani Nenkova, James Thomas, Byron C Wallace

**Affiliations:** School of Population Health and Environmental Sciences, King’s College London, London, UK; Khoury College of Computer Sciences, Northeastern University, Boston, US; Vortext Systems, Groningen, Netherlands; Cochrane Dementia group, University of Oxford, Oxford, UK; Cochrane Editorial Unit, London, UK; Computer and Information Science, University of Pennsylvania, Philadelphia, US; EPPI-Centre, UCL, London, UK

## Abstract

**Objective:** Randomized controlled trials (RCTs) are the gold standard method for evaluating whether a treatment works in healthcare, but can be difficult to find and make use of. We describe the development and evaluation of a system to automatically find and categorize all new RCT reports.

**Materials and Methods:** *Trialstreamer*, continuously monitors PubMed and the WHO International Clinical Trials Registry Platform (ICTRP), looking for new RCTs in humans using a validated classifier. We combine machine learning and rule-based methods to extract information from the RCT abstracts, including free-text descriptions of trial populations, interventions and outcomes (the ‘PICO’) and map these snippets to normalised MeSH vocabulary terms. We additionally identify sample sizes, predict the risk of bias, and extract text conveying key findings. We store all extracted data in a database which we make freely available for download, and via a search portal, which allows users to enter structured clinical queries. Results are ranked automatically to prioritize larger and higher-quality studies.

**Results:** As of May 2020, we have indexed 669,895 publications of RCTs, of which 18,485 were published in the first four months of 2020 (144/day). We additionally include 303,319 trial registrations from ICTRP. The median trial sample size in the RCTs was 66.

**Conclusions:** We present an automated system for finding and categorising RCTs. This yields a novel resource: A database of structured information automatically extracted for all published RCTs in humans. We make daily updates of this database available on our website (https://trialstreamer.robotreviewer.net).

## Background and Significance

Randomized controlled trials (RCTs) are the gold standard study design to determine what works in health, [1] and access to the results of such trials is central to the practice of evidence-based medicine. However, finding and making use of RCT evidence can be difficult [2]. The number of RCTs published accelerates every year,[3] but they still make up a tiny fraction of the contents of health research databases.[4] It is therefore difficult to retrieve trials relevant to particular clinical questions.

An up-to-date resource comprising all published RCTs and data on ongoing trials would facilitate efficient retrieval of the best-available evidence, especially if it allowed for structured search with respect to individual PICO elements (i.e., study Populations, Interventions/Comparators, and Outcomes).[5] This would be a boon to healthcare practitioners and to researchers interested in seeking the latest evidence for a given question at the point of care, or in gaining a broad overview of all clinical evidence on particular topics, e.g., as in a *scoping review* or similar evidence mapping exercise.

In this paper we describe our development of such a system, which we call *Trialstreamer*. Using a validated machine learning model,[4] Trialstreamer identifies reports of new RCTs in humans as they are published and runs these through a suite of trained data extraction models to extract elements of interest, including: study sample sizes, key findings, descriptions of the PICO elements, and an indicator of the risk of bias.

We store all identified RCT reports and associated extracted data in a publicly available, continuously updated database. We believe this will be a valuable resource to the larger informatics community. We also make the data accessible via an open-source prototype web-interface (Figures 2 and 3; https://trialstreamer.robotreviewer.net/). This interface capitalizes on the extracted data to allow users to precisely structure queries to address a clinical question of interest, and to automatically rank retrieved articles to prioritise the largest and highest quality trials.

**Figure 1.**
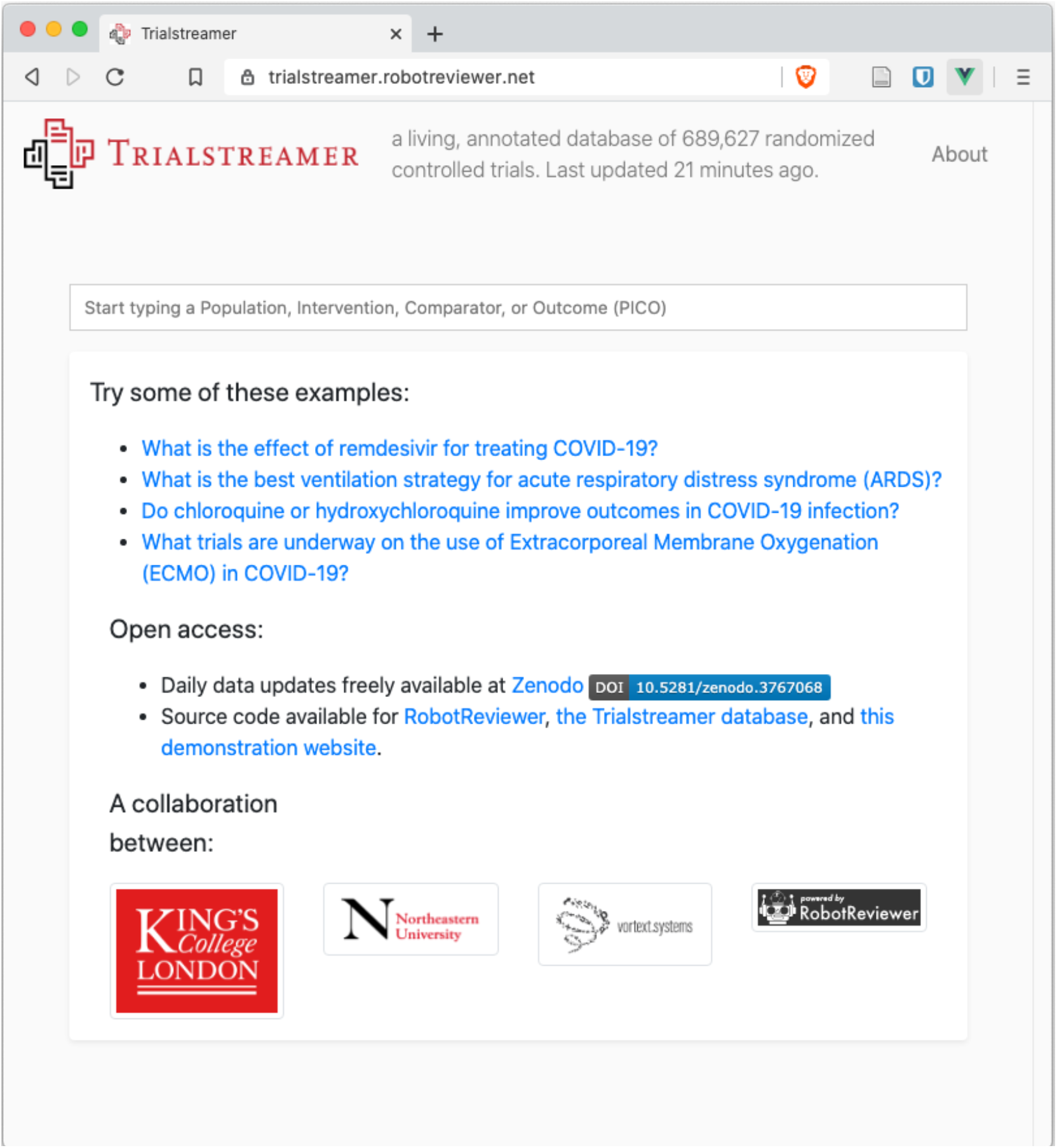
Screenshot of the Trialstreamer web interface home page

**Figure 2.**
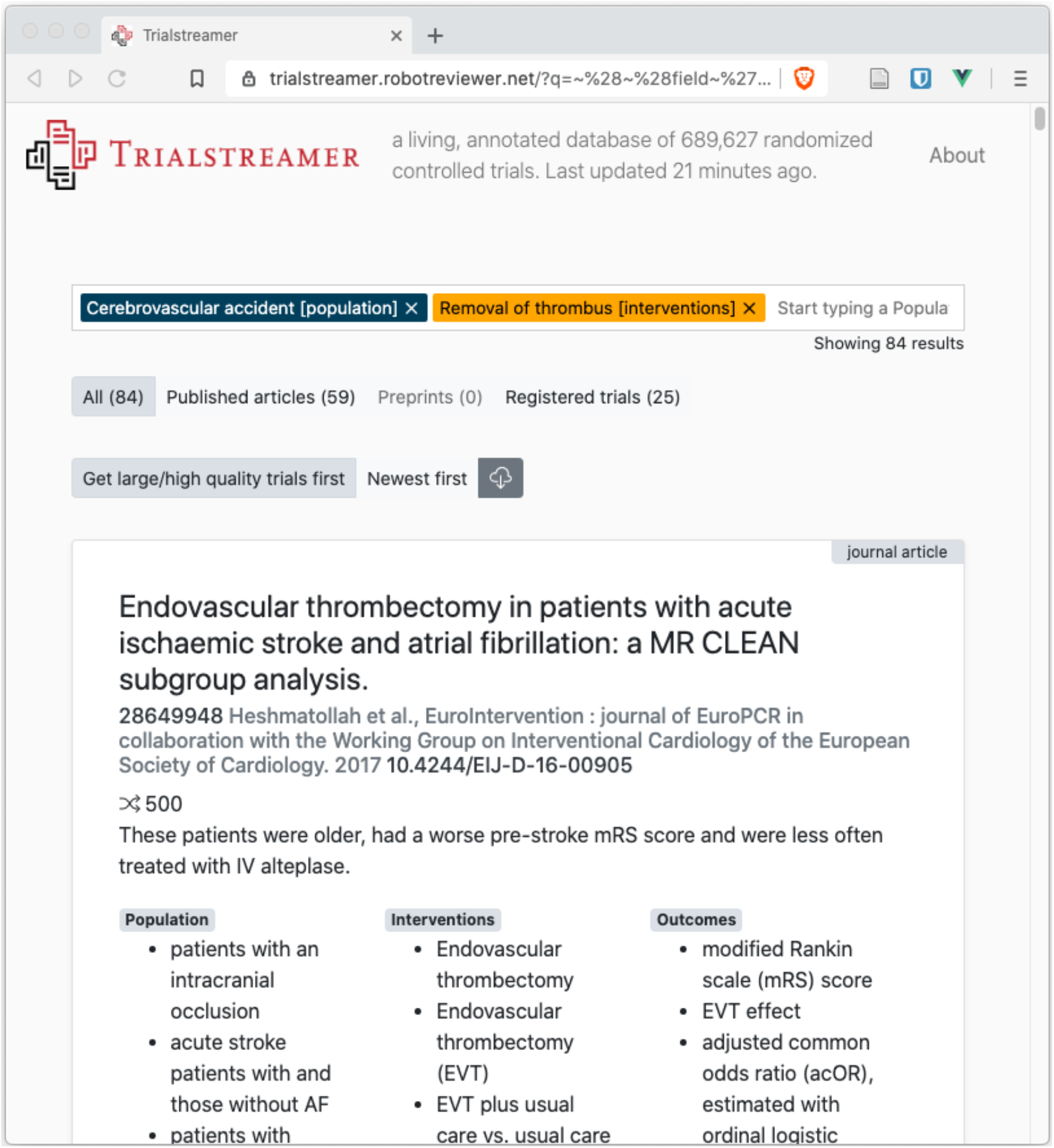
Trialstreamer search results

**Figure 3.**
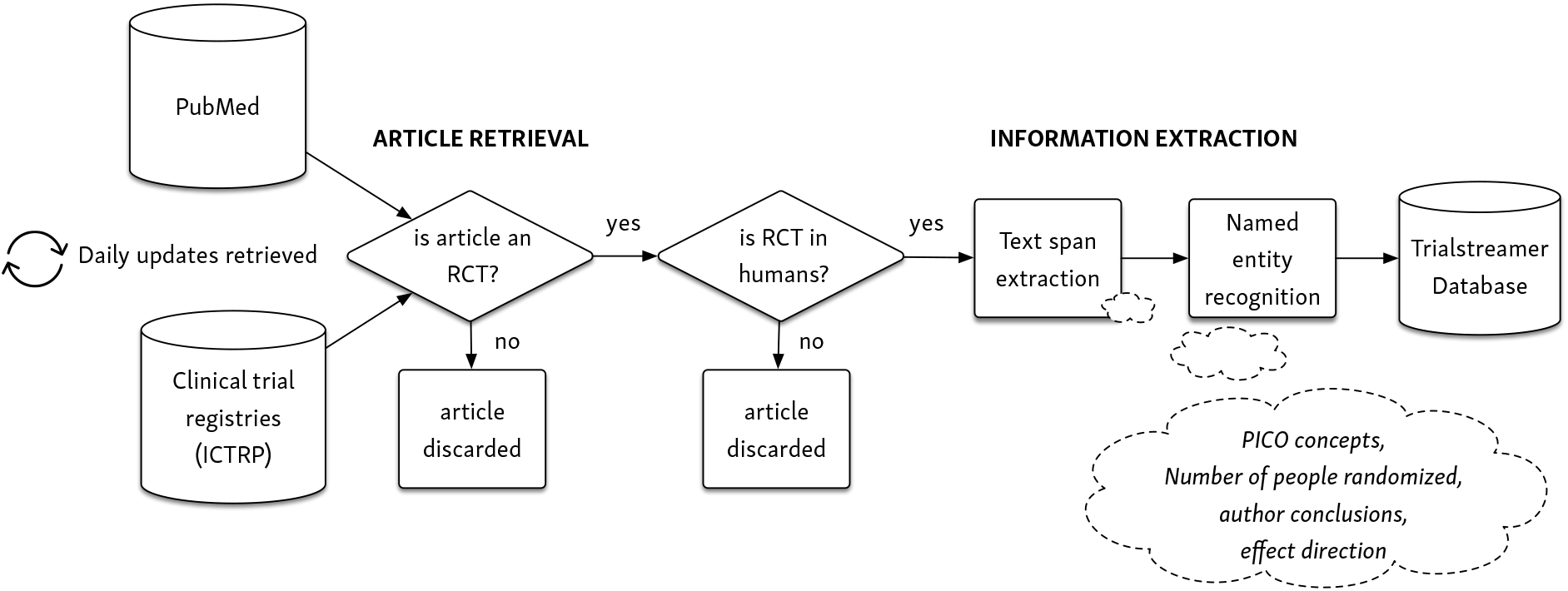
How articles are retrieved, annotated, and stored

## Objective

We describe the Trialstreamer database, the elements it comprises, and how we keep the database up-to-date. We review the machine learning models we use to extract individual elements from trial reports, and we empirically evaluate the accuracy of the system components. We make the data freely accessible in several ways: via our prototype web portal, bulk data download from the open science platform, Zenodo (https://doi.org/10.5281/zenodo.3767068), or by executing the Trialstreamer source code locally to reproduce the database.

## Materials and Methods

### OVERVIEW

Trialstreamer monitors key research databases, seeking to retrieve reports of RCTs in humans quickly after publication. The architecture of the system is summarised in Figure 3 and Table 1. First, we collect peer-reviewed journal articles (via PubMed), and registrations of ongoing trials (via the WHO International Clinical Trials Registry Platform [ICTRP]).^1^ From these sources we retrieve *all* research articles and study registrations. We then use a machine learning system to determine the study design of each article. Articles classified as RCTs in humans are retained, and other study designs are discarded. This step should discard > 95% of the source articles. Next, key data are extracted from the RCT abstracts (namely a description of the PICO, the number of participants, a snippet stating the main findings, and an assessment of the risk of bias). The articles together with the extracted information items are saved to a relational database. The full process is quick: articles are typically available in the database (with full annotations) less than an hour after release by PubMed. The updated database is accessible via a web search portal, and also for bulk download. We provide an overview of the machine learning systems for each step below. All code for generating the database and the machine learning models is freely available via our website^2^. We describe each step in detail below.

**Table 1.** Summary of Trialstreamer components

| Component | Model architecture | How used in Trialstreamer | Data |
| --- | --- | --- | --- |
| RCT vs non-RCT study classifier | Ensemble of Support Vector Machine and Convolutional Neural Network models[4] | Previously validated model[4] used with new 'balanced' classification threshold | Training and parameter tuning on 280,000 abstracts with crowdsourced labels from Cochrane Crowd<br>Evaluation on 49,028 abstracts manually labelled by the Clinical Hedges team |
| Human vs non-human study classifier | Support vector machine model, based on Cohen *et al.*[6] | New model trained and validated, based on prior method | 467,153 abstracts of RCTs from PubMed, using MeSH term "Humans"<br>Training and parameter tuning on 75% of data, evaluation on withheld 25% of data |
| Sample size extraction | Multilayer perceptron model for classifying integers in abstracts | New model trained and validated | Trained on 8,935 abstracts with sample sizes (6,315 taken from structured data in ClinicalTrials.gov; 2620 manually labelled by our team)<br>Evaluation on 500 separate abstracts manually labelled by the authors |
| PICO text spans | LSTM-CRF model[7] | Previously validated model used unchanged[7] | 5,000 abstracts with from *EBM-NLP* dataset[7]<br>Model training/parameter tuning on 4,800 abstracts labeled by lay crowdworkers.<br>Evaluation on 200 withheld abstracts labeled by medical doctors |
| PICO concepts | Rule-based concept extraction, following the method of *Metamap Lite* by Demner-Fushman *et al*[8] | Reimplementation of previously described method | 972,371 selected concepts (CUIs) and their associated text from the UMLS Metathesaurus 2019 edition. Concepts included were from the source vocabularies: SNOMED CT, RxNorm, MeSH, MedDRA [Medical Dictionary for Regulatory Activities], and the World Health Organization ATC [Anatomic Therapeutic Chemical] classification system<br>Evaluation on 1,497 unseen abstracts with crowd-sourced PICO MeSH labels from Cochrane Crowd |
| Risk of bias | Logistic regression model with L2 regularization, using bag-of-words text representation (unigrams, bigrams and trigrams) to generate an overall score | New model trained and validated | 13,463 abstracts of RCTs with Cochrane Risk of Bias tool assessments in the Cochrane Library.<br>60% used for training; 40% withheld for evaluation |

### ARTICLE RETRIEVAL

#### Identifying RCTs

PubMed indexes > 30 million research articles^3^, and is growing rapidly. We are interested in the subset of these articles that describe RCTs, and more specifically RCTs conducted in humans. Such articles comprise 1–2% of the total.[4] While we could use the high-quality manual indexes applied by PubMed staff (namely, the ‘Randomized Controlled Trial’ Publication Type, and the ‘Humans’ MeSH^4^ term), there is several months’ lag from publication date to indexing, given that this is a manual process. This would result in our system failing to include recently published material.

Instead, we use a machine learning classifier that we have described previously to continuously identify new RCT reports.[4] In short, this system uses an ensemble of models comprising a Support Vector Machine (SVM), and ten independent Convolutional Neural Network (CNN) models. These models were trained on a set of 280,000 abstracts manually labeled by the *Cochrane Crowd* project.^5^ For articles that have a manually applied Publication Type, we incorporate this information as an additional input feature for the ensemble.

Our prior work focused on classifying RCTs for systematic review searches; such reviews emphasize finding *all* relevant studies. Therefore, for that application we designed the classifier to have very high recall (near 99%), which as a trade-off corresponded to relatively low precision (≈20%). The assumption here is that the model serves as an initial filter, and that the output would then go through a manual screening step, such that the remaining irrelevant articles would be excluded.

By contrast, Trialstreamer seeks to be useful for clinical question answering and literature scoping; it is intended to be used fully automatically, without a manual screening step. We therefore calibrated and evaluated a new classification threshold that seeks a more appropriate balance of recall and precision for this use. To achieve this, we drew Receiver Operating Characteristics (ROC) curves for the trained models with regard to a set of 49,028 abstracts labeled by the McMaster University Clinical Hedges team.[9] We set a threshold which maximised the sum of sensitivity and specificity (i.e. the outer left extreme of the curve). To evaluate the performance of this strategy, we conducted 5000 bootstrap iterations, where a classification threshold was set on a random sample with replacement of the Hedges dataset, and evaluated on the data not included in that sample.[10] We evaluated the accuracy of binary prediction (precision/recall) and calibration (via calibration curves, Brier scores, and the cstatistic). On the same validation dataset, we previously found that the manual PubMed Publication Type index had recall 0.94, and precision 0.56 for retrieving RCTs.[4]

#### Identifying studies conducted in humans

We next remove any RCTs not conducted in humans (e.g., animal or agricultural studies) using an SVM model, following the method described by Cohen and colleagues.[6] Here, we trained a similar model using abstracts of RCTs from PubMed, with labels derived as a function of whether the MeSH term ‘Humans’ had been applied or not. We used 350,364 abstracts for training, and evaluated performance on 116,789 withheld abstracts (25% of the dataset), and evaluated using 1000 bootstrap iterations. We add all studies determined to be both RCTs and in humans to the database; these go forward for (automated) annotation.

### AUTOMATED ANNOTATION OF RCTs

#### Sample Size Extraction

We assume that if the sample size is reported in the abstract, it will be as an integer (either in numerals or words). We first use a series of heuristics and regular expressions to convert numbers expressed in natural language to numerals (e.g., “one hundred and twelve” would be replaced with “112”). We then use a Multilayer Perceptron (MLP) model to estimate the probability that each integer represents the study sample size. This model uses a set of features describing the integer context in the abstract. These include word embeddings of adjacent tokens (initialized to weights from a *word2vec* model[11] pre-trained over a large corpus of PubMed articles[12]), inferred part-of-speech tags for surrounding words, and bespoke features that indicate, e.g., whether some variant of the word “patients” occurs near the token under consideration.

The model was trained using 8,935 RCT abstracts for which the true number of people randomized was available: 6,315 of these labels were derived automatically using registry data from ClinicalTrials.gov; the remaining 2,620 were labelled manually by members of our team. We provide the complete feature set and other model details in the Appendix.

#### PICO Extraction

The penultimate step in our pipeline entails extracting snippets from abstracts that describe the Population enrolled, the Interventions and Controls administered, and the Outcomes evaluated (the PICO elements) in the trial being described. We then automatically map these snippets to concept unique MeSH identifiers.

To extract the text snippets, we use a modelling approach we have described in detail previously,[7] and summarise here. We use a Long Short Term Memory (LSTM)-Conditional Random Field (CRF) — i.e., an LSTM-CRF[13] — to make token-level predictions.

In this model, input texts (represented as token and character-level embeddings) are passed through an LSTM layer to yield contextualized representations of words. The token embeddings were initialized using pre-trained word vectors.[12] These representations are then passed through a CRF layer, which makes predictions as to whether each word is a Population, Intervention/Comparator, or Outcome. We trained the model on the ∼5,000 RCT abstracts that comprise the “EBM-NLP” dataset.[7] This dataset contains abstracts with manually annotated text snippets describing each element of the PICO.

As a second step we expand any abbreviations within the extracted text snippets by algorithmically identifying abbreviation definitions (e.g. “Atrial Fibrillation (AF)”) which appear elsewhere in the text.[14]

Finally, we follow the method set out by Demner-Fuchman and colleagues, who created the software ‘MetaMap Lite’.^14^ Briefly, a database of synonyms for medical terms is automatically compiled by retrieving all the free-text descriptions of the concepts included in vocabularies contained in the UMLS Metathesaurus. These synonyms are minimally pre-processed (including, for example, removing database tags such as ‘[NOS]’, and lowercasing all terms). The end result is an index of synonyms for each MeSH term. These synonyms can be sought in the extracted text snippet describing the trial population to find associated MeSH terms. We use a different set of vocabularies compared with MetaMap Lite to generate our synonyms (namely SNOMED CT, RxNorm, MeSH, MedDRA [Medical Dictionary for Regulatory Activities], and the World Health Organization ATC [Anatomic Therapeutic Chemical] classification system). These are also the vocabularies chosen by the Cochrane linked data project,^6^ specifically designed to describe PICO attributes from clinical trial reports in Cochrane Reviews, and we found that they gave good coverage for PICO concepts during development. We evaluate these PICO concept extraction steps against 1,437 abstracts which had been manually labelled by the Cochrane Crowd project with structured PICO terms. Our main evaluation assesses whether the system produces exact matches to the manual terms (which we label the ‘strict’ evaluation).

The UMLS Metathesauraus encodes conceptual relationships between terms (for example, noting “Anterior Wall Myocardial Infarction” is related to “Myocardial Infarction”, the latter being the parent term). Given adjacent terms are often sufficiently close in meaning for practical purposes, we also report a ‘relaxed’ evaluation, which in addition to exact matches, considers immediate parent and child terms as correct.

#### Risk of bias

A key task in evidence-based medicine is determining whether problems in study methods might bias the results. We have previously described in detail methods for automatically assessing the risk of bias in full text RCTs (in PDF form) for the purposes of conducting a systematic review.[15–17] In this prior work, we predicted whether studies’ results might be at risk of bias using the first version of the Cochrane Risk of Bias tool^7^ (which examines whether bias is likely due to problems in the random sequence generation, allocation concealment, and blinding, among other issues).[18]

For the Trialstreamer database, we adjust our approach in two ways. First, we consider abstracts only (using 57,144 abstracts of RCTs which had been manually assessed using the Cochrane Risk of Bias tool in systematic reviews in the Cochrane Library). In related work, Millard and colleagues found that predicting risks of bias from abstracts was possible, though modestly less accurately than using full-texts.[19] Second, here we seek to use risk of bias predictions as an input to inform search rankings, as compared with our prior work in which we aimed to produce a definitive prediction for each bias domain for use in a systematic review (after human review).

For Trialstreamer, making predictions for particular subcategories of bias, (say, asserting that the random sequence was adequately generated) is likely to be misleading to the user, given that this information is usually not explicitly in the abstract. Instead, we prefer to generate a score for an *overall* risk of bias, using the rule of thumb outlined in the Cochrane Handbook.[20] Using this approach, studies are rated as being at ‘low’ risk of bias overall, when *all* the subcategories assessed are rated ‘low’. If one or more subcategories are rated high or unclear, the overall risk of bias is rated high or unclear.

Since our goal was to produce well-calibrated scores, we use a logistic regression model. The article title and abstract represented in bag-of-words form with unigram, bigram, and trigram features. We used L2 regularization.[21]

We trained this model using 51,429 abstracts and their associated manual ‘overall’ bias assessments as labels; we evaluated the model on 5,715 withheld articles. We calculated model performance both in terms of binary predictive accuracy (precision, recall/sensitivity, and specificity) and calibration accuracy (via a calibration curve, Brier score, c-statistic, and shrinkage parameters). All metrics with confidence intervals were estimated using bootstrap resampling (using 1,000 iterations).[10]

#### The Prototype Web Portal

We make the annotated data accessible for browsing via a web portal (https://trialstreamer.robotreviewer.net; screenshot in Figures 1 and 2). The search portal features an autocompleter, which suggests structured terms (in Population, Interventions, or Outcomes facets) when the user begins typing a query. Articles matching the query are automatically retrieved; for each of these we present the data elements that our models have extracted (screenshot in Figure 4). The user may rank the search retrieval by sample size, and by risk of bias; thus allowing the largest and highest quality research to be prioritised. The portal also allows filtering by published trials, preprints, and registered (i.e. ongoing) trials. Retrieved articles can be exported in the RIS format for use in citation managers.

**Figure 4.**
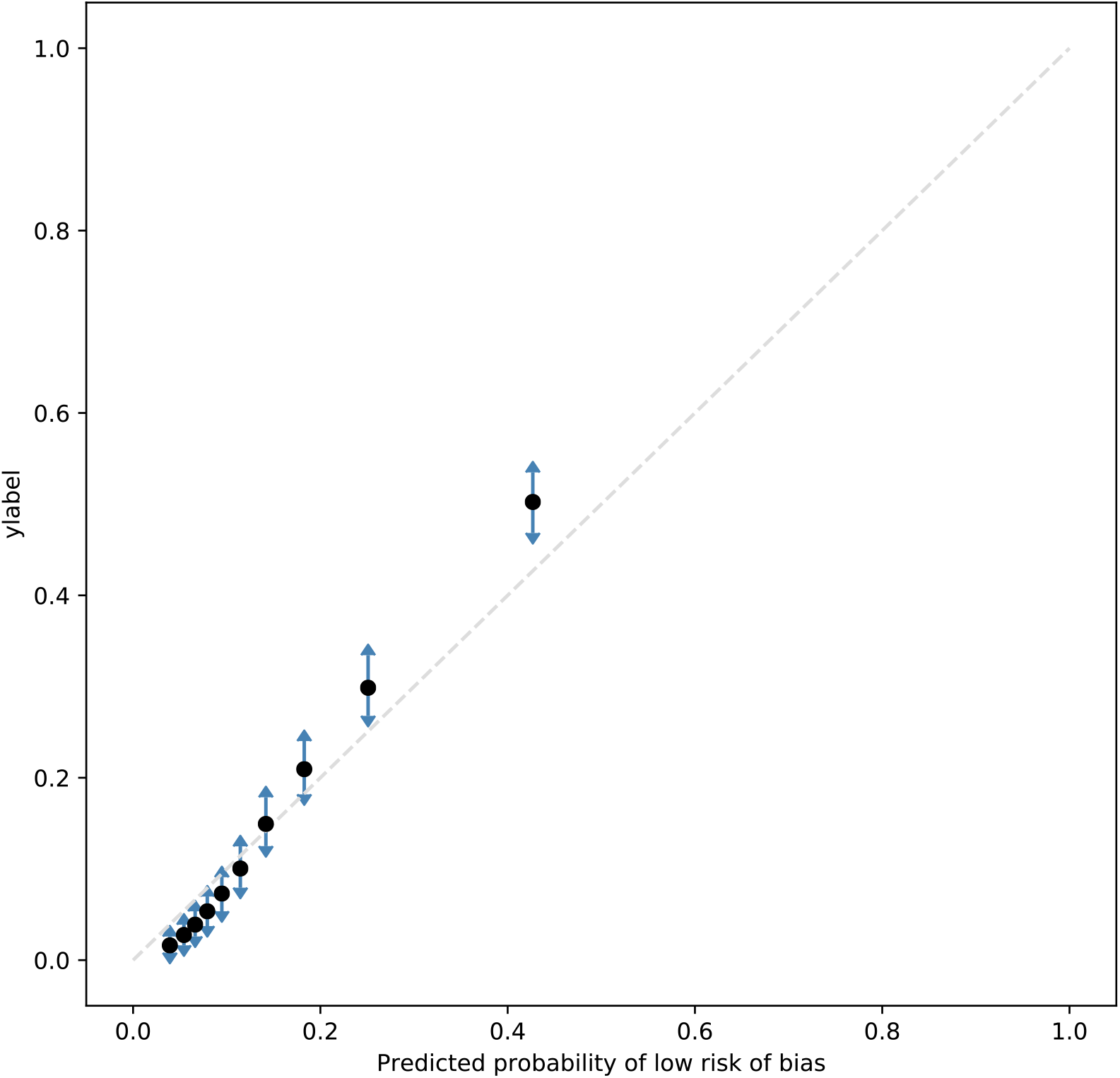
Calibration plot of Risk of Bias model

### Results

#### Validation of system performance

We summarize the performance of the system components in Table 2. The RCT classifier retrieved 94–97% of RCT articles, with the search retrieval realizing ≥50% precision; the human study classifier had near perfect precision and recall.

**Table 2.** Summary of evaluation performance of models used in the Trialstreamer system. *The PICO text span accuracy results are taken from Nye et al.,[7] and we present here for convenience

| Component | Recall | Precision | C statistic | Brier score |
| --- | --- | --- | --- | --- |
| RCT classifier (balanced threshold) |  |  |  |  |
| MeSH indexed articles | 0.97 (0.96, 0.98) | 0.52 (0.48, 0.56) | 0.99 (0.98, 0.99) | 0.01 (0.01, 0.01) |
| Not MeSH indexed | 0.94 (0.92, 0.96) | 0.50 (0.44, 0.57) | 0.98 (0.98, 0.98) | 0.01 (0.01, 0.01) |
| Human vs non-human classifier | 1.00 (1.00, 1.00) | 1.00 (1.00, 1.00) | 0.95 (0.94, 0.96) | 0.003 (0.003, 0.004) |
| Sample size extraction | 0.79 (0.77, 0.82) | 0.88 (0.85, 0.90) | n/a | n/a |
| PICO text spans* |  |  |  |  |
| Population (n=200) | 0.66 | 0.78 | n/a | n/a |
| Interventions (n=200) | 0.65 | 0.61 |  |  |
| Outcomes (n=200) | 0.63 | 0.69 |  |  |
| PICO concepts<br>*strict* |  |  |  |  |
| Population (n=1107) | 0.73, (0.70, 0.75) | 0.28, (0.26, 0.29) | n/a | n/a |
| Interventions (n=954) | 0.78, (0.75, 0.80) | 0.52, (0.49, 0.55) |  |  |
| Outcomes (n=503) | 0.64, (0.61, 0.68) | 0.25, (0.23, 0.27) |  |  |
| *Relaxed* |  |  |  |  |
| Population (n=1107) | 0.78, (0.76, 0.81) | 0.30, (0.29, 0.32) |  |  |
| Interventions (n=954) | 0.85, (0.82, 0.87) | 0.57, (0.54, 0.60) |  |  |
| Outcomes (n=503) | 0.65, (0.62, 0.69) | 0.30, (0.27, 0.33) |  |  |
| Risk of bias (probability of being at low risk of bias) | 0.46, (0.40, 0.52) | 0.44, (0.41, 0.48) | 0.80, (0.79, 0.81) | 0.10, (0.10, 0.11) |

We use the model for identifying PICO spans directly from Nye and colleagues.[7] This evaluation found F1 scores of 0.71, 0.65, and 0.63 for tagging individual words as describing Populations, Interventions/Comparators, and Outcomes, respectively. These are per-word (or “token”) metrics, and are therefore pessimistic; extracted snippets that do not perfectly align with reference span annotations may still be high-quality, and anecdotally we find this is often the case.

We found that MeSH labels for PICO concepts had good recall for populations and interventions (recall of 0.78 and 0.85 respectively), but lower precision (0.28 and 0.52). On qualitative evaluation, we found that many of the apparent errors in precision could be regarded as correct, and were due to the high specificity of the Cochrane Crowd dataset used in the evaluation (where typically only the most important concept was labelled in a study). For example, the most common false positive terms in our evaluation for Population were ‘Patients’ (8%) and ‘Elderly’ (3%); the most common false positive Intervention labels were ‘Placebo’ (10%) and ‘Therapeutic procedure’ (3%). We therefore conducted a manual re-evaluation of the precision for Population and Interventions labels, where one author not involved in the system development (R. Maclean, a medical doctor) re-assessed a random sample of 100 abstracts with their predicted labels. This evaluation found a precision of 0.79 for Population descriptors, and 0.84 for Intervention descriptors.

The model for predicting whether an RCT was at low risk of bias showed good calibration and ranking ability (Brier score 0.10, c-statistic=0.80, and see calibration plot in Figure 4). Binary classification was less accurate with regard to the gold standard labels (F1=0.45).

#### Database contents

On 8th May 2020, there were 669,895 RCTs indexed in the Trialstreamer database, of which 18,485 had been published in the first four months of 2020 (compared with 968 manually indexed in MEDLINE at the same time point). This equates to 144 RCTs published daily in 2020 to date. We additionally include 303,319 records from We compare the manual indexing in PubMed versus the automated system in Figure 5. We present the distribution of sample sizes in the trials in Figure 6. The median sample size was 66, with interquartile range 30 to 181).

**Figure 5.**
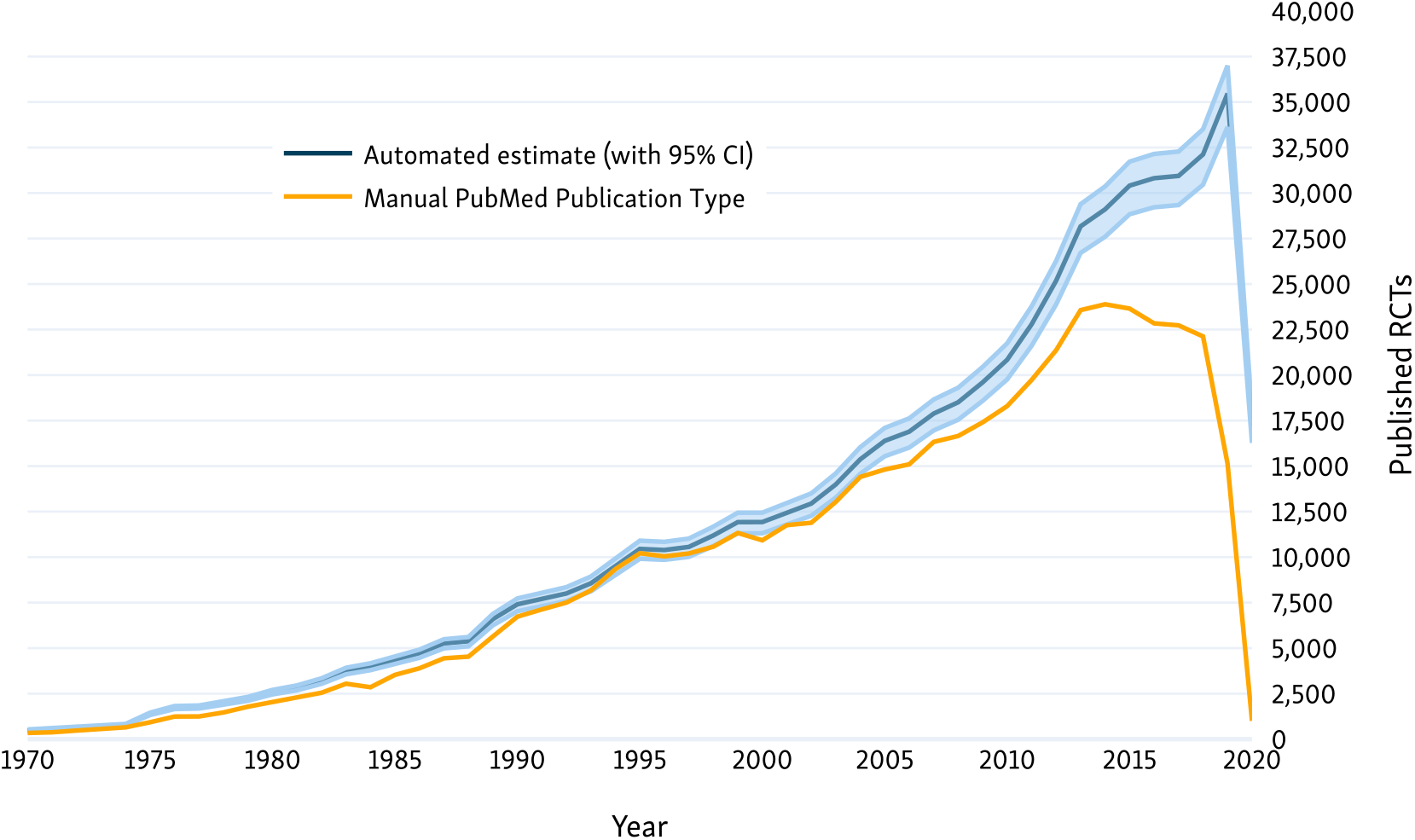
Counts of all RCTs in PubMed, estimated by manual indexing (yellow) versus automation (blue)

**Figure 6.**
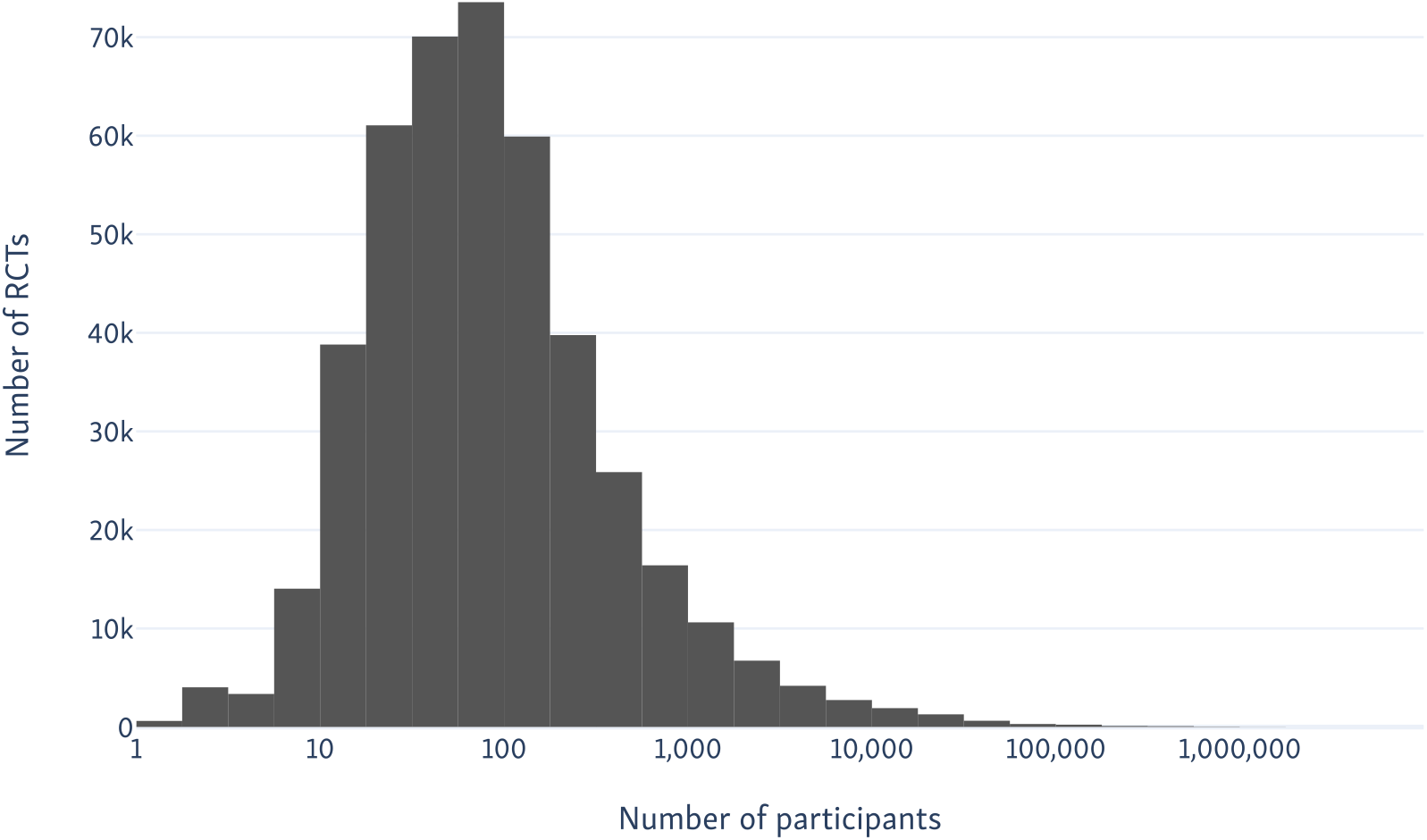
Histogram of the number of trial participants in all RCTs in PubMed, as extracted by our sample size extraction model.

### Discussion

At present, accessing all evidence reported in RCTs in humans relevant to a particular query or clinical question is difficult for a few key reasons. First, if one relies on manual indexing of RCTs (e.g., relying on the Publication Type tag in PubMed), this will necessarily constitute an incomplete subset due to the lag inherent to manual categorization (Figure 5). Second, trials are reported in unstructured free-text, which means that retrieving those relevant to a specific question (or PICO frame) is non-trivial. Further, even once relevant trials are retrieved, one must manually appraise the quality of trials, extract sample sizes, and peruse the text to infer what the reported results are. All of this takes time, a problem exacerbated by the rapid expansion of the biomedical literature.

Trialstreamer uses machine learning and natural language processing to address these issues, in turn providing a new publicly available resource which we make available to the broader community: A continuously updated, comprehensive database of all published RCTs in humans, with semi-structured data extracted for all of them. This resource may facilitate additional research (e.g., allowing one to investigate the clinical topics for which there exists a paucity of RCT evidence), evidence scoping (or “rapid reviews”), and clinical question answering for healthcare practitioners interested in efficiently seeking out new trial results related to a specific question.

No machine learning models have perfect accuracy, and we have made some pragmatic decisions around the use of models, which have strengths and limitations. Our system recalls 97% of RCTs (as compared with the standard used in systematic review search filters, which achieve ≈98.5% recall[4]). However, we trade off this small fraction of missed articles for substantially higher precision which should allow the data to be used for real-time question answering or scoping, without the need for extensive manual filtering. In future, we plan to evaluate the use effectiveness of the system for rapid answering of clinical questions relating to the effectiveness of healthcare interventions.

Likewise, our model for assessing the risk of bias has on face value, poor performance for binary classification (given recall of 0.45, a substantial portion of high quality studies would be missed in a binary classification mode). However, the model probabilities show good calibration, and thus are useful in the current system as a means of ranking documents, and surfacing the highest quality evidence first.

### Conclusions

We have introduced Trialstreamer, a living, continuously updated database of semi-structured information automatically extracted from all published clinical trials as they are published. We provided quantitative evaluations of all individual components comprising Trialstreamer. We make both the underlying database available, as well as a public web interface that facilitates search. Additionally, all code (including model implementations, database creation scripts, and our prototype) for Trialstreamer is open-source.

## Data Availability

We release freely our code, models, and the Trialstreamer database on our website, and via the Zenodo open science platform (https://doi.org/10.5281/zenodo.3767068). The EBM-NLM dataset is available freely online. This work makes use of data created by Cochrane, Cochrane Crowd, and Clinical Hedges at MacMaster University, and the Unified Medial Language System (UMLS) from the National Library of Medicine, which are available directly from the owners, subject to the copyright holder’s terms.

https://trialstreamer.robotreviewer.net

https://doi.org/10.5281/zenodo.3767068

## Author contributions

Conceived the study: IJM & BCW, with input from JT, R Marshall, AN and FS. Creation of training and evaluation data: ANS, JT, BN, IJM, AN, and BCW. Coding of machine learning models: IJM, BN, and BCW. Manual model evaluation R Maclean. Coding of web portal: IJM, JK, and BCW. Initial draft of manuscript: IJM & BCW. Critical revisions of manuscript, and sign-off of final version: all.

## Acknowledgements

We would like to thank the Cochrane Crowd team and the volunteer annotators, who produced the training data for RCT classifcation and evaluating our PICO models. We also acknowledge Cochane for generous provision of systematic review data for research purposes, which we used to create the system for assessing bias in RCTs. We are enormously grateful to Clinical Hedges team at the Health Information Research Unit at McMaster University for making their data available for our evaluation.

## Footnotes

1 During the current COVID-19 pandemic, we additionally monitor preprints of COVID-19 trials from medRxiv.

2 https://trialstreamer.robotreviewer.net

3 https://pubmed.ncbi.nlm.nih.gov/

4 MeSH refers to the Medical Subject Headings vocabulary, maintained by the National Library of Medicine (NLM) and used for indexing articles in the MEDLINE database.

5 https://crowd.cochrane.org/

6 https://linkeddata.cochrane.org/

7 We note that a second version of the Risk of Bias tool has been released. We stick with the first version owing to the availability of training data.

## References

1 Chalmers I. The Cochrane Collaboration: Preparing, Maintaining, and Disseminating Systematic Reviews of the Effects of Health Care. Ann N Y Acad Sci 1993;703:156–65. doi:10.1111/j.1749-6632.1993.tb26345.x

2 Shaughnessy AF, Slawson DC, Bennett JH. Becoming an information master: a guidebook to the medical information jungle. J Fam Pr 1994;39:489–99.

3 Bastian H, Glasziou P, Chalmers I. Seventy-Five Trials and Eleven Systematic Reviews a Day: How Will We Ever Keep Up? PLOS Med 2010;7:e1000326. doi:10.1371/journal.pmed.1000326

4 Marshall IJ, Noel-Storr A, Kuiper J, et al. Machine learning for identifying Randomized Controlled Trials: An evaluation and practitioner’s guide. Res Synth Methods Published Online First: 4 January 2018. doi:10.1002/jrsm.1287

5 Thomas J, Kneale D, McKenzie JE, et al. Chapter 2: Determining the scope of the review and the questions it will address. In: Cochrane Handbook for Systematic Reviews of Interventions. Cochrane 2019. www.training.cochrane.org/handbook

6 Cohen AM, Dunivin ZO, Smalheiser NR. A probabilistic automated tagger to identify human-related publications. Database J Biol Databases Curation 2018;2018. doi:10.1093/database/bay079

7 Nye B, Jessy Li J, Patel R, et al. A Corpus with Multi-Level Annotations of Patients, Interventions and Outcomes to Support Language Processing for Medical Literature. ACL 2018;2018:197–207.

8 Demner-Fushman D, Rogers WJ, Aronson AR. MetaMap Lite: an evaluation of a new Java implementation of MetaMap. J Am Med Inform Assoc 2017;24:841–4. doi:10.1093/jamia/ocw177

9 Montori VM, Wilczynski NL, Morgan D, et al. Optimal search strategies for retrieving systematic reviews from Medline: analytical survey. BMJ 2005;330:68. doi:10.1136/bmj.38336.804167.47

10 Steyerberg EW, Harrell FE Jr, Borsboom GJ, et al. Internal validation of predictive models: efficiency of some procedures. J Clin Epidemiol 2001;54:774–81. doi:10.1016/S0895-4356(01)00341-9

11 Mikolov T, Sutskever I, Chen K, et al. Distributed Representations of Words and Phrases and their Compositionality. In: Burges CJC, Bottou L, Welling M, et al., eds. Advances in Neural Information Processing Systems 26. Curran Associates, Inc. 2013. 3111–3119. http://papers.nips.cc/paper/5021-distributed-representations-of-words-and-phrases-and-their-compositionality.pdf (accessed 12 May 2020).

12 Pyysalo S, Ginter F, Moen H, et al. Distributional semantics resources for biomedical text processing. Proc Lang Biol Med 2013.

13 Huang Z, Xu W, Yu K. Bidirectional LSTM-CRF Models for Sequence Tagging. ArXiv150801991 Cs Published Online First: 9 August 2015. http://arxiv.org/abs/1508.01991 (accessed 11 May 2020).

14 Schwartz AS, Hearst MA. A simple algorithm for identifying abbreviation definitions in biomedical text. Pac Symp Biocomput Pac Symp Biocomput 2003;:451–62.

15 Marshall IJ, Kuiper J, Wallace BC. Automating risk of bias assessment for clinical trials. 2014. 88–95.

16 Marshall IJ, Kuiper J, Wallace BC. RobotReviewer: evaluation of a system for automatically assessing bias in clinical trials. JAMIA 2016;23:193–201. doi:10.1093/jamia/ocv044

17 Zhang Y, Marshall I, Wallace BC. Rationale-Augmented Convolutional Neural Networks for Text Classification. In: EMNLP. 2016. 795–804. https://www.ncbi.nlm.nih.gov/pubmed/28191551

18 Higgins JPT, Altman DG, Gotzsche PC, et al. The Cochrane Collaboration’s tool for assessing risk of bias in randomised trials. BMJ 2011;343:d5928–d5928. doi:10.1136/bmj.d5928

19 Millard LA, Flach PA, Higgins JP. Machine learning to assist risk-of-bias assessments in systematic reviews. Int J Epidemiol 2016;45:266–77. doi:10.1093/ije/dyv306

20 Higgins JPT, Deeks JJ. Chapter 7: Selecting studies and collecting data. In: Cochrane Handbook for Systematic Reviews of Interventions. John Wiley & Sons 2008.7.1–7.28.

21 Ng AY. Feature selection, L 1 vs. L 2 regularization, and rotational invariance. Proc Twenty-First Int Conf… 2004;:78.

22 Kingma DP, Ba J. Adam: A Method for Stochastic Optimization. ArXiv14126980 Cs Published Online First: 29 January 2017. http://arxiv.org/abs/1412.6980 (accessed 24 Apr 2020).

